# Kidney Transplant Recipients and Omicron: Outcomes, effect of vaccines and the efficacy and safety of novel treatments

**DOI:** 10.1101/2022.05.03.22274524

**Authors:** Sarah Gleeson, Paul Martin, Tina Thomson, Amarpreet Thind, Maria Prendecki, Katrina J. Spensley, Candice L. Clarke, Candice Roufosse, Graham Pickard, David Thomas, Stephen P. McAdoo, Liz Lightstone, Peter Kelleher, Michelle Willicombe

## Abstract

We aimed to describe the outcomes of Omicron infection in kidney transplant recipients (KTR), compare the efficacy of the community therapeutic interventions and report the safety profile of molnupiravir.

From 142 KTRs diagnosed with COVID-19 infection after Omicron had become the dominant variant in the UK, 116 (78.9%) cases were diagnosed in the community; 47 receiving sotrovimab, 21 molnupiravir and 48 no treatment. 10 (20.8%) non-treated patients were hospitalised following infection, which was significantly higher than the sotrovimab group (2.1%), p=0.0048, but not the molnupiravir treated group (14.3%), p=0.47. The only admission following sotrovimab occurred in a patient infected with BA.2. One patient from the molnupiravir and no-treatment groups required ICU support, and both subsequently died, with one other death in the no-treatment group. No patient receiving sotrovimab died. 6/116 (5.2%) patients required dialysis following their diagnosis; 2 (9.5%) patients receiving molnupiravir and 4 (8.3%) no-treatment. This requirement was significantly higher in the molnupiravir group compared with the sotrovimab treated patients, in whom no patient required dialysis, p=0.035. Both molnupiravir treated patients requiring dialysis had features of systemic thrombotic microangiopathy.

Post-vaccination serostatus was available in 110 patients. Seropositive patients were less likely to require hospital admission compared with seronegative patients, 6 (7.0%) and 6 (25.0%) respectively, p=0.023. Seropositive patients were also less likely to require dialysis therapy, p=0.016.

In conclusion, sotrovimab treatment in the community was associated with superior patient and transplant outcomes; it’s clinical efficacy against the BA.2 variant requires a rapid review. The treatment benefit of molnupiravir was not evident, and wider safety reporting in transplant patients is needed.

## Introduction

Whilst Omicron has been considered the least pathogenic COVID-19 variant in the general population, infection in solid organ transplant recipients cannot be assumed to be ‘mild’^1,2^. With diminished protection from vaccination compared with the general population, prompt treatment following diagnosis now offers further protection against an adverse outcome in this vulnerable population^3,4^. In the UK, community therapeutic options for people at high risk of deterioration included intravenous sotrovimab (xevudy) or oral molnupiravir (lagevrio) from 21^st^ December 2021, with oral nirmatrelvir plus ritonavir (paxlovid) and intravenous remdesivir (veklury) being introduced from 10^th^ February 2022^5^.

The studies which informed the use of sotrovimab and molnupiravir have important caveats when applied to transplant recipients in the Omicron era. The efficacy of sotrovimab, a human monoclonal antibody that neutralizes SARS-CoV-2, was demonstrated in the alpha variant era prior to vaccination, although, reassuringly, sotrovimab has continued to show neutralising properties against BA.1, in *in vitro* studies ^6^. The study also excluded transplant recipients, although it is likely that the results are applicable to immunosuppressed populations and, it could be hypothesised that the use of neutralising antibodies in immune suppressed individuals may be more beneficial than in the immunocompetent.^6^. Molnupiravir, an oral anti-viral drug, modified from a compound known as NHC (β-d-N4 -hydroxycytidine), was shown to be efficacious in preventing hospitalisation in people considered high risk in a study which ran from October 2020. However, people with renal impairment, defined as a glomerular filtration rate (GFR) ≤30mls/min were excluded^7^. Although not cleared by the kidneys, with negligible amounts detected in urine, neither it’s safety or efficacy had been tested in patients with significant chronic kidney disease (CKD) prior to its use^8^. Molnupiravir was granted conditional approval for use in the UK and the US, but not in Europe and some other countries^9^.

This study aims to describe the outcomes of Omicron infection in kidney transplant recipients (KTR), compare the efficacy of the different therapeutic interventions and report the safety profile of molnupiravir in KTRs.

### Methods

We prospectively identified transplant recipients diagnosed with COVID-19 infection between 17^th^ December 2021, at which time Omicron infection was the dominant variant in London, until 31^st^ March 2022. All patients were under the care of Imperial College Renal and Transplant Centre, London. Clinical and pathology data were obtained from electronic patient records. The study was approved by the Health Research Authority, Research Ethics Committee (Reference: 20/WA/0123 - The Impact of COVID-19 on Patients with Renal disease and Immunosuppressed Patients).

### COVID-19 Management

In the UK, transplant outpatients with COVID-19 infection were able to access therapeutics via COVID medicines delivery units (CMDU) following referral from the health care team. Treatment was offered if patients fulfilled the following criteria: (i) infection confirmed via reverse-transcriptase polymerase chain reaction (RT-PCR) testing, (ii) symptomatic disease, and (iii) would start therapy within 5 days of testing. Treatment options from 21^st^ December 2021-10^th^ February 2022 were either a 5-day course of the oral anti-viral molnupiravir or a single 500mg infusion of xevudy (sotrovimab). Patients were offered the choice of treatment.

From 10^th^ February 2022, the inclusion criterion expanded to include infections diagnosed via lateral flow antigen testing. Therapeutics offered were also extended to include the anti-viral paxlovid (nirmatrelvir/ritonavir) or sotrovimab as first line options. With intravenous remdesivir and molnupiravir as 2^nd^- and 3^rd^ line respectively.

In addition to community treatment, when the transplant centre was notified, we would recommend that patients receiving anti-proliferative agents stop them for a 14-day period.

### UK Vaccination schedule

Transplant patients were offered either BNT162b2 (Pfizer) or ChAdOx1 (AZ) as 1^st^ and 2nd COVID doses in the UK. In response to data showing inadequate vaccine immune responses in immunosuppressed people, a 3^rd^ primary dose of a mRNA vaccine was offered from September 2021. The UK responses to the emergence of the Omicron variant precipitated a booster dose for all of the population, including transplant recipients, from December 2021. All booster doses were mRNA vaccines.

### Diagnosis and genotyping

For the duration of the study, RT-PCR and lateral flow testing was free to access in the UK and all transplant patients were provided with a home nasopharyngeal swab kit for RT-PCR testing to use as required. Genotyping was not routinely performed on RT-PCR specimens from outpatients. RT-PCR samples from patients admitted to hospital were used for sequencing until the Omicron variant accounted for 100% of positive cases in London, after which time Omicron infection was diagnosed using S-gene target failure (SGTF).

### SARS-CoV-2 antibody detection

Routine serological testing of transplant patients has implemented in our NHS trust since June 2020. For vaccine responses, anti-S IgG were assessed using the Abbott Architect SARS-CoV-2 IgG Quant II CMIA. Anti-S antibody titres are quantitative with a threshold value of 7.1 BAU/ml for positivity, and an upper level of detection of 5680 BAU/ml.

### Statistical Analysis

Statistical analysis was conducted using MedCalc v20.106. Unless otherwise stated, all data are reported as median with interquartile range (IQR). The Chi-squared test was used for proportional assessments. Using the log rank test, Kaplan-Meier analyses were used to estimate and compare clinical outcomes by community therapeutic interventions.

## Results

During the study period, 142 KTRs were diagnosed with COVID-19 infection. Five (3.5%) patients had confirmed Delta-variant infection, 15 (10.6%) were diagnosed with Omicron infection at or during an in-patient admission and the remaining 122 (85.6%) were diagnosed in the community with presumed Omicron infection, **Figure 1**. Clinical details of the patients infected by the Delta-variant maybe found in the *Supplemental Information*, **Table S1**.

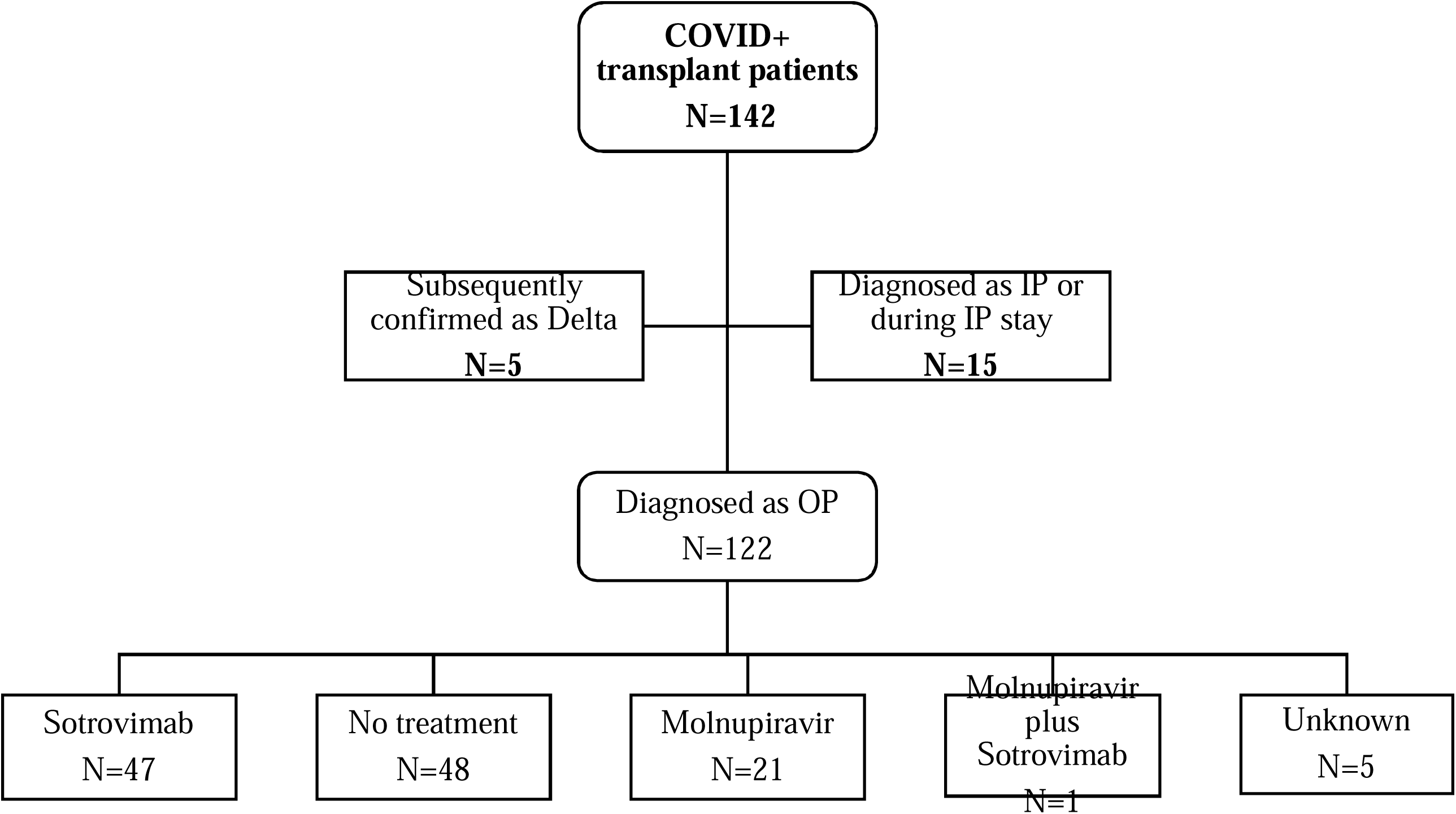

Management interventions for the 122 outpatients (OP) consisted of sotrovimab in 47 (38.5%) cases, molnupiravir in 21 (17.2%), no treatment in 48 (39.3%), molnupiravir plus sotrovimab in 1 patient and data was missing in 5 cases, **Figure 1**. The latter 6 cases were excluded from further analysis, and the remaining 116 patients were followed up for a median of 97 (76-110) days post diagnosis.

### Comparison of demographic features by treatment

Compared with patients who received sotrovimab, patients receiving molnupiravir were less likely to be Caucasian (p=0.001) or have received ≥3 doses of vaccine (p=0.0038). No other significant differences were seen between the 2 groups, **Table 1**. Patients receiving ‘no-treatment’ were more likely to have received less than 3 doses of vaccines compared with the sotrovimab treated group (p=0.0046), **Table 1**. The ‘no-treatment’ group were also more likely to have a diagnosis of diabetes (p=0.043), and have reached end stage kidney disease (ESKD) secondary to APKD (autosomal dominant polycystic kidney disease) (p=0.013), compared with the sotrovimab treated patients.

### Patient and allograft outcomes following a community COVID-19 diagnosis

There was no statistical difference in the incidence of hospitalisation in patients who received molnupiravir compared with sotrovimab, 3 (14.3%) and 1 (2.1%) of patients in each group respectively (p=0.056) **Figure 2**. Sequencing of the patient admitted following sotrovimab administration confirmed an Omicron BA.2 infection, which was the only case of the BA.2 sublineage detected in the OP setting in this analysis. Ten of 48 (20.8%) non-treated patients were hospitalised following an infection diagnosis, which was significantly higher than the sotrovimab group (p=0.0048) but not the molnupiravir treated group (p=0.47), **Figure 2**. One patient from each of the molnupiravir and no-treatment groups required ICU for respiratory support for COVID pneumonitis.

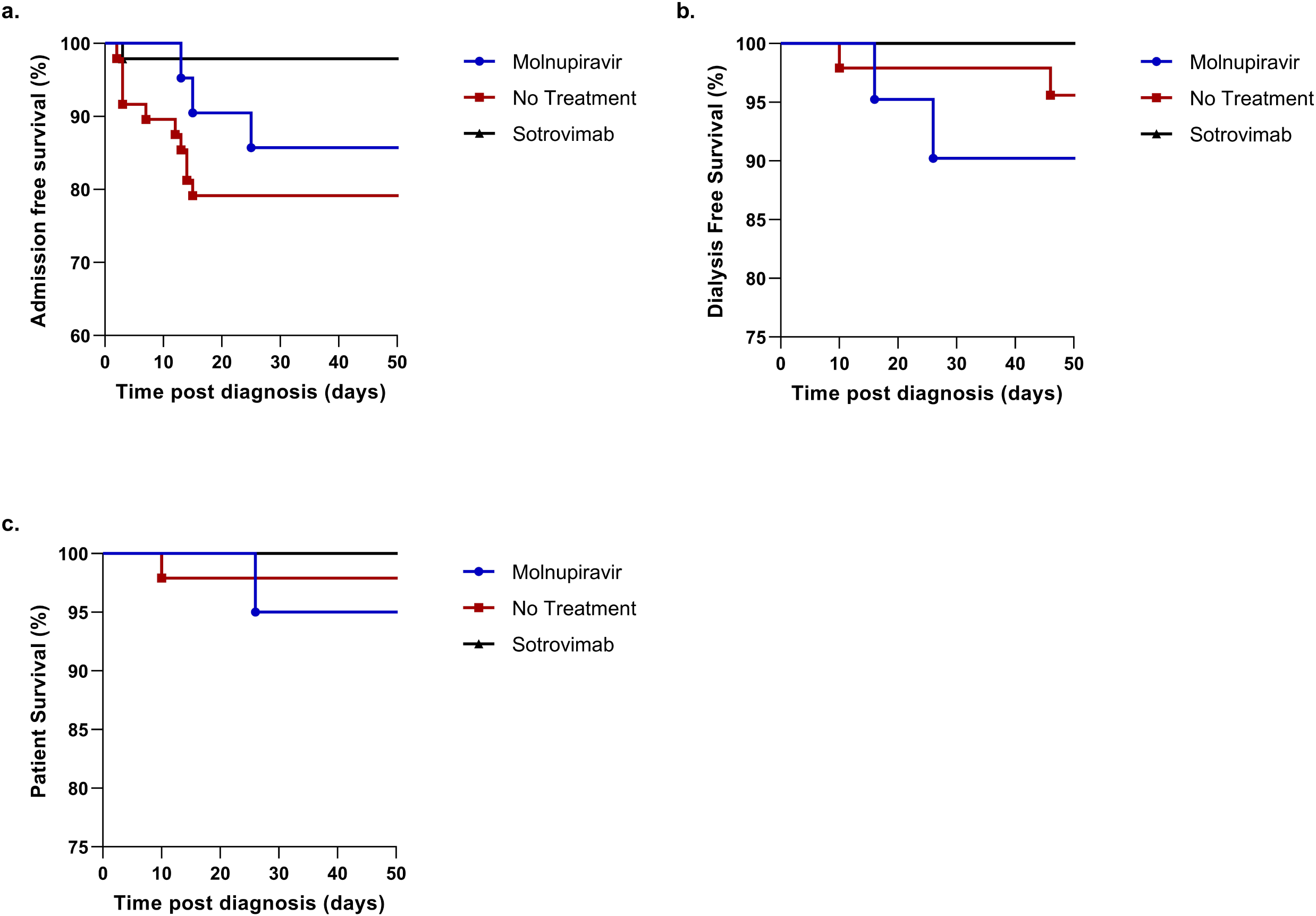

Six of 116 (5.2%) patients required either dialysis or haemofiltration following their diagnosis; 2 (9.5%) patients receiving molnupiravir and 4 (8.3%) no-treatment. This requirement was significantly higher in the molnupiravir group compared with the sotrovimab treated patients, in whom no patient required dialysis, (p=0.035), **Figure 2**. Both molnupiravir treated patients requiring acute renal support presented with features of systemic thrombotic microangiopathy (TMA). One of these patients, had a renal allograft biopsy showing arteriolar and glomerular thrombi consistent with TMA in addition to pyelonephritis. Two other molnupiravir treated patients have undergone an allograft biopsy; one with acute graft dysfunction post infection whose biopsy showed T cell rejection and one (biopsy planned pre infection for gradually worsening graft function) whose biopsy showed acute glomerular ischemia and a likely organising thrombus in an arteriole consistent with TMA. Four patients in the no treatment group have had a biopsy post infection, none of which showed any features of TMA. There was no significant difference in the need for renal support between the groups who received sotrovimab compared with no-treatment (p=0.069).

Three patients (2.6%) have subsequently died; 2 who received no-treatment, including one death due to COVID-19 infection, and another COVID-19 death in a patient who had received molnupiravir. No deaths were recorded in a patient who received sotrovimab, **Figure 2**.

### Patient and allograft outcomes following a community diagnosis of COVID-19 infection by serostatus

Serostatus post vaccination was available in 110 patients: post 2-doses in 38 patients, post 3-doses in 59 patients and 4-doses in 13 patients. Twenty-four (22%) patients were ‘non-responders’, and 86 (78%) responders, with a median anti-S concentration of 522 (102-1449) BAU/ml. Seronegative patients were more likely to require hospital admission following infection compared with seropositive patients; 6 (25.0%) and 6 (7.0%) of non-responders and responders respectively (p=0.023), **Figure 3**. Six patients in each cohort required admission. Seronegative patients were also more likely to require dialysis support; 3 (12.5%) and 1 (1.2%) of non-responders and responders respectively (p=0.016), **Figure 3**. During the follow up period, there was no difference in patient survival between the groups, with 1 and 2 deaths in the seropositive and seronegative groups respectively. The anti-S concentration in the seropositive patient who died was 103 BAU/ml, which was measured 31 days prior to infection.

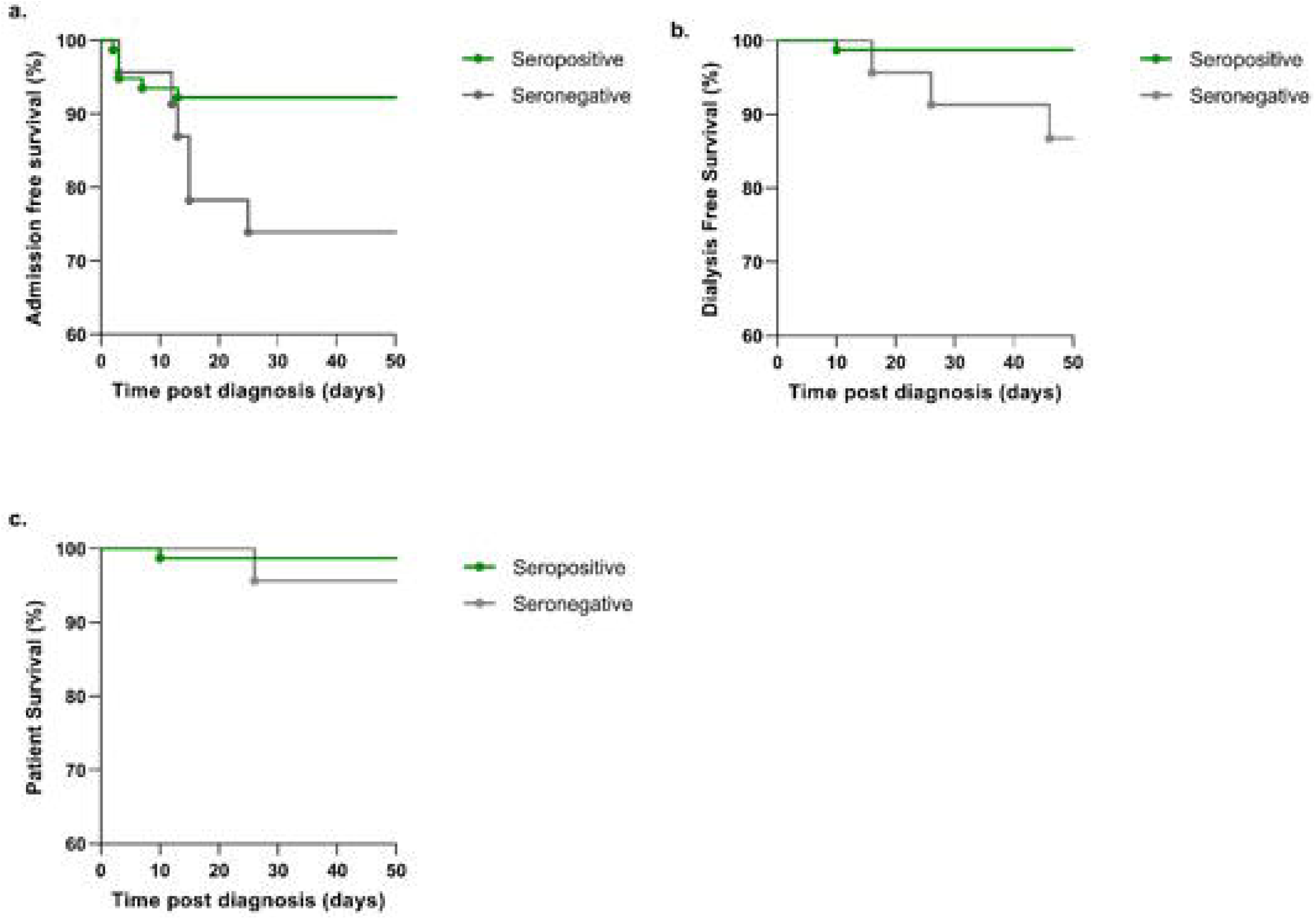

### Patient and allograft outcomes following an in-patient COVID-19 diagnosis

The clinical demographics and treatment of the patients diagnosed with Omicron infection in the inpatient setting maybe found in the *Supplemental Information*, **Table S2**. Of the 15 patients diagnosed, 7 (46.7%) were positive at the time of presentation to hospital, whilst 8 (53.3%) were considered to have acquired nosocomial infection. Only 2/7 (28.6%) of those diagnosed at presentation were considered to have COVID as their primary diagnosis. Both required acute haemodialysis, with one cared for on the ICU, and dying 46 days following admission. Six of the 8 patients with nosocomial infection received treatment, with 1 patient acutely deteriorating prior to administration and 1 patient not requiring treatment.

### Discussion

Within the study limitations, we have shown that sotrovimab treatment in the community is associated with a reduction in hospital admission compared with KTRs who received no-treatment or molnupiravir. In addition, sotrovimab was shown to lead to superior transplant outcomes compared with KTRs who received molnupiravir. However, sotrovimab treated patients were also more likely to have completed their primary vaccine course with or without boosters, which suggests differing baseline populations.

In addition to efficacy reporting, this study also raises a potential safety concern for the use of molnupiravir in KTRs. Although we report no difference in the need for dialysis support in patients receiving molnupiravir versus no-treatment, the presence of clinical features compatible with thrombotic microangiopathy (TMA) in the molnupiravir group warrants urgent attention. Thrombotic microangiopathy is not uncommon in KTRs, but its occurrence with the addition of systemic features are rare^10^. Viral infections and drugs are both recognised causes of TMA in KTRs, but the lack of TMA in the no-treatment or sotrovimab groups may suggest a treatment precipitant, either due to its direct actions or interactions with concomitant medications. Also of note, the baseline renal function of the molnupiravir treated patients with adverse outcome was less than 30ml/min of GFR, that is below the inclusion criteria for the molnupiravir ‘MOVe-OUT’ study^7^.

In this study, we have also demonstrated the importance of continued screening and genotyping of immunocompromised individuals whilst the pandemic is ongoing. The reason for this is twofold. Firstly, at a population level, it is recognised that evolution of the virus may occur in the immunocompromised host^11^. Secondly, our study shows how viral sequencing may influence the management of the infected KTRs. Sotrovimab has been shown to have less efficacy against the BA.2 variant, and it was recognised that our only patient admitted post sotrovimab treatment had sequencing confirming BA.2 infection. Genotyping of this individual at the time of diagnosis could have led to alternative management^12^. However, there are limited alternative treatment options currently in the UK with only molnupiravir, paxlovid or remdesivir available. Given our findings above, we would be reluctant to use molnupiravir, given the multiple drug interactions, paxlovid is not an attractive option for KTRs, whilst remdesivir is contraindicated for those with a GFR of less than 30mls/min, which precludes a significant proportion of KTRs as well as being logistically challenging to deliver. Therefore, KTRs remain in an unfavourable predicament, being in a group with poorer outcomes following infection, suboptimal protection from vaccination, and limited treatment options^3^. Alternative prophylaxis, with evusheld, a combination of 2 monoclonal antibodies tixagevimab and cilgavimab, may be of benefit for this vulnerable population in the UK, and is already being used elsewhere in the world^13^. However, it is also recognised that these monoclonal antibodies may not stay effecttive against infection as the virus continues to evolve, and other strategies should also be developed^13^.

It may be considered that even within a KTR population that there remains a spectrum of risk in terms of infection outcome following vaccination. Perhaps, alternative prophylaxis could be reserved for those at highest risk? We found that serostatus associates with clinical outcome, and mechanistically this makes sense, as non-responder status is likely to identify those KTRs who have the highest level of immune suppression. Our group has previously provided our opinion on the use of antibody testing post-vaccination for this very purpose, and although we recognise the caveats of such a proposal, it could be a pragmatic way to deliver personalised medicine for this group of patients with greatest need^14^.

This descriptive observational study has small numbers which preclude definitive conclusions in an unselected patient cohort with differing baseline characteristics. However, it highlights the need for transplant specific research into the optimal COVID-19 interventions. Although recognised as a high-risk group with poor infection outcomes, immunosuppressed people often with significant comorbidities, have been excluded from policy informing studies from the start of the pandemic. Relying on extrapolating data from these studies to inform management of transplant recipients, may and has led to suboptimal outcomes.

## Data Availability

all relevant data in manuscript

## Acknowledgments

This research is supported by the National Institute for Health Research Biomedical Research Center based at the Imperial College Healthcare NHS Trust and Imperial College London. The authors thank the West London Kidney Patient Association and all the patients and staff at ICHNT (the Imperial COVID vaccine group and dialysis staff and staff within the North West London Pathology laboratories). The authors are also grateful for support from The Nan Diamond Fund, Sidharth and Indira Burman, and the Auchi Charitable Foundation. SG is supported by a Kidney Research UK Clinical Research Fellowship. MP is supported by a National Institute for Health Research clinical lectureship. KJS is supported by a National Institute for Health Research Academic Clinical Fellowship. Work in DT’s laboratory is supported by a Wellcome Trust Clinical Career Development Fellowship.

