## Supplementary material for "Kidney Transplant Recipients and Omicron: Outcomes, effect of vaccines and the efficacy and safety of novel treatments": Table 1

**Table 1. Demographics of 116 patients diagnosed with COVID-19 infection in the community**

| Characteristics | | Sotrovimab  N=47 (%) | Molnupiravir  N=21 (%) | No Treatment  N=48 (%) | P value |
| --- | --- | --- | --- | --- | --- |
| Gender | Male  Female | 22 (46.8)  25 (53.2) | 10 (47.6)  11 (52.4) | 31 (64.6)  17 (35.4) | 0.18 |
| Age at 1^st^ vaccine | Years (Median) | 53 (42-64) | 44 (37-57) | 55 (48-66) | 0.10 |
| Ethnicity | Caucasian  Black  Indoasian  Other | 27 (57.4)  -  17 (36.2)  3 (6.4) | 3 (14.3)  1 (4.8)  12 (57.1)  5 (23.8) | 19 (39.6)  3 (6.2)  19 (39.6)  7 (14.6) | 0.024 |
| Cause of ESKD | Polycystic kidney disease  Glomerulonephritis  Diabetic nephropathy  Urological  Unknown  Other | 8 (17.0)  18 (38.3)  5 (10.6)  1 (2.1)  6 (12.8)  9 (19.1) | -  9 (42.9)  4 (19.0)  2 (9.5)  4 (19.0)  2 (9.5) | 1 (2.1)  14 (29.2)  12 (25.0)  6 (12.5)  9 (18.8)  6 (12.5) | 0.06 |
| Number of transplants received | 1  ≥2 | 43 (91.5)  4 (8.5) | 21 (100.0)  - | 46 (95.8)  2 (4.2) | 0.31 |
| 1^st^ vaccine pre-transplant | No  Yes | 41 (87.2)  6 (12.8) | 21 (100.0)  - | 39 (81.2)  9 (18.8) | 0.10 |
| Diagnosis in 1^st^ year post -tx | No  Yes | 41 (87.2)  6 (12.8) | 20 (95.2)  1 (4.8) | 39 (81.2)  9 (18.8) | 0.29 |
| Type of transplant | Deceased Donor  Living Donor  Simultaneous Pancreas-Kidney | 24 (51.1)  21 (44.7)  2 (4.3) | 12 (57.1)  7 (33.3)  2 (9.5) | 31 (64.6)  21 (44.7)  2 (4.3) | 0.45 |
| Induction agent | Alemtuzumab  IL2 receptor antagonist  None  Unknown | 33 (70.2)  3 (6.4)  11 (23.4) | 15 (71.4)  3 (14.3)  3 (14.3) | 29 (60.4)  7 (14.6)  12 (25.0) | 0.58 |
| Immunosuppression type | CNI Monotherapy  CNI/MMF (orAza)  CNI/MMF/Prednisolone  CNI/Prednisolone  Other | 18 (38.3)  15 (31.9)  10 (21.3)  4 (8.5)  - | 9 (42.9)  4 (19.0)  6 (28.6)  2 (9.5)  - | 22 (45.8)  8 (16.7)  10 (20.8)  6 (12.5)  2 (4.2) | 0.77 |
| Diabetes | No  Yes | 39 (83.0)  8 (17.0) | 14 (66.7)  7 (33.3) | 31 (64.6)  17 (35.4) | 0.11 |
| No. of vaccines | 0  1  2  3  4 | -  -  1 (2.1)  28 (59.6)  18 (38.3) | 2 (9.5)  -  3 (14.3)  14 (66.7)  2 (9.5) | -  1 (2.1)  9 (18.8)  33 (68.7)  5 (10.4) | 0.0007 |
| Anti-S protein pre-infection | BAU (Median) | 136 (24-1230) | 175 (10-682) | 108 (7.1-677) | 0.58 |
| Seronegative | No  Yes  Missing | 37 (78.7)  10 (21.3) | 16 (76.2)  5 (23.8) | 34 (70.8)  12 (25.0)  2 (4.2) | 0.53 |
| Baseline function | Creatinine (mmol/L)  eGFR (ml/min/1.73m2) | 109 (84-159(  57 (35-72) | 127 (98-190)  48 (29-73) | 118 (94-160)  55 (35-72) | 0.54 |
