## Supplemental information for "Kidney Transplant Recipients and Omicron: Outcomes, effect of vaccines and the efficacy and safety of novel treatments"

### **Table S1. Characteristics of hospitalised patients presenting with Delta-variant infections during the study period**

| **Age** | **GENDER** | **ETHNICITY** | **IMMUNO-**  **SUPPRESSION** | **DIABETES** | **# OF VACCINES** | **ANTI-S** | **ICU** | **COMMUNITY TREATMENT** | **IP TREATMENT** | **RIP** |
| --- | --- | --- | --- | --- | --- | --- | --- | --- | --- | --- |
| In 50s | MALE | WHITE | FK/MMF | NO | 3 | 9.46 | YES | NIL | DEX/REM/RONAPREVE | YES |
| In 50s | MALE | WHITE | FK/MMF | NO | 3 | 7.1 | YES | MOLNUPIRAVIR | DEX/RONAPREVE | NO |
| In 50s | FEMALE | WHITE | FK/MMF | YES | 3 | 7.1 | YES | NIL | DEX/REM/RONAPREVE/SOTROVIMAB | NO |
| In 50s | FEMALE | WHITE | FK/MMF | YES | 3 | 7.1 | YES | NIL | UNKNOWN | NO |
| In 50s | MALE | WHITE | FK/MMF/PRED | NO | 3 | 7.1 | YES | NIL | DEX/REM/RONAPREVE | NO |

### **Table S2. Characteristics of patients diagnosed with Omicron-variant infections at presentation or during IP stay during the study period**

| **Age** | **GENDER** | **ETHNICITY** | **IMMUNO-**  **SUPPRESSION** | **DIABETES** | **# OF VACCINES** | **ANTI-S** | **Diagnosis** | **IP TREATMENT** | **ICU** | **RIP** |
| --- | --- | --- | --- | --- | --- | --- | --- | --- | --- | --- |
| In 60s | Female | WHITE | FK | YES | 3 | 222 | During inpatient stay | NO TREATMENT | YES | YES |
| In 60s | Female | Asian | FK | NO | 3 | 3864 | During inpatient stay | SOTROVIMAB | NO | NO |
| In 50s | Male | WHITE | FK/MMF | YES | 3 | 740 | During inpatient stay | SOTROVIMAB | YES | NO |
| In 70s | Female | OTHER | FK | YES | 3 | 505 | During inpatient stay | TOCI/DEX | NO | NO |
| In 70s | Female | ASIAN | FK/MMF | NO | 3 | 7.1 | During inpatient stay | NO TREATMENT | NO | NO |
| In 60s | Male | ASIAN | FK | YES | 3 | 49 | During inpatient stay | SOTROVIMAB | NO | NO |
| In 60s | Male | BLACK | FK | YES | 0 | 7.1 | At presentation | INCIDENTAL | NO | NO |
| In 50s | Male | WHITE | FK | NO | 0 | 55 | During inpatient stay | SOTROVIMAB | NO | NO |
| In 60s | Male | WHITE | FK | YES | 2 | 7.1 | During inpatient stay | SOTROVIMAB | NO | NO |
| In 70s | Male | ASIAN | FK | YES | 3 | 12.7 | At presentation | INCIDENTAL | NO | NO |
| In 70s | Female | ASIAN | FK | NO | 3 | 7.1 | At presentation | NO TREATMENT | NO | NO |
| In 40s | Male | WHITE | FK | NO | 2 | 18.0 | At presentation | NO TREATMENT | NO | NO |
| In 50s | Female | OTHER | FK/MMF/PRED | NO | 2 | 7.1 | At presentation | REMDES | NO | NO |
| In 60s | Male | ASIAN | FK/MMF | NO | 4 | 7.1 | At presentation | SOT/TOCI/REMES/DEX | NO | NO |
| In 50s | Female | BLACK | FK/MMF/PRED | YES | 2 | 18 | At presentation | TOCI/DEX | YES | NO |
